# Geometry-Aware Reproducibility of Imputation Protocol (GRIP): diagnosing two failure modes of single imputation for heavy-tailed, collinear variables in biomedical data

**DOI:** 10.64898/2026.07.14.26358017

**Authors:** Claire Su-Yeon Park

**Author notes:** Corresponding author: /.

## Abstract

**Background:** Imputation accuracy is typically summarized by a single figure computed from one set of random deletions. For the heavy-tailed, collinear variables common in biomedical data, this study asks whether that figure can be trusted, and introduces a diagnostic protocol that determines when it cannot.

**Methods:** This paper introduces GRIP (Geometry-aware Reproducibility of Imputation Protocol), a three-step diagnostic that profiles each variable’s geometry and collinearity, stress-tests reproducibility under MCAR, MAR, and MNAR missingness with fixed seeds, and classifies the failure mode. GRIP was demonstrated on 1,885 United States Centers for Medicare & Medicaid Services (CMS) home-health agencies (15 numeric variables), comparing missForest, votingForest, mean, and median imputation. A supplementary k-nearest-neighbour (k-NN) grid (20 bivariate lognormal parameter combinations; n = 500) verified the SILENT failure boundary across imputer types. The simulation component followed the ADEMP framework.

**Results:** Geometry profiling prospectively flagged two variables combining extreme right-skew (skewness 38.6, 43.3) with near-collinearity (|r| = 0.96). Under MCAR and MAR, missForest normalized root mean squared error (NRMSE) ranged from below 1 to 152 across otherwise identical replications (SD 14–30); the difficulty ordering of the two variables reversed in 69% of replications. Under MNAR self-masking, instability vanished (SD = 0), yet only 14% of true extreme magnitudes were recovered. Both failure modes arise from one mechanism: instability requires an extreme value to be absent while its collinear partner remains observed. A k-NN proxy grid confirmed SILENT failure in 11 of 15 high-skew parameter combinations under MNAR, regardless of correlation level.

**Conclusions:** For heavy-tailed, collinear variables, one imputation-accuracy number can mislead in two opposite ways. GRIP detects both before an imputer is committed and is provided as reusable, open-source R code.

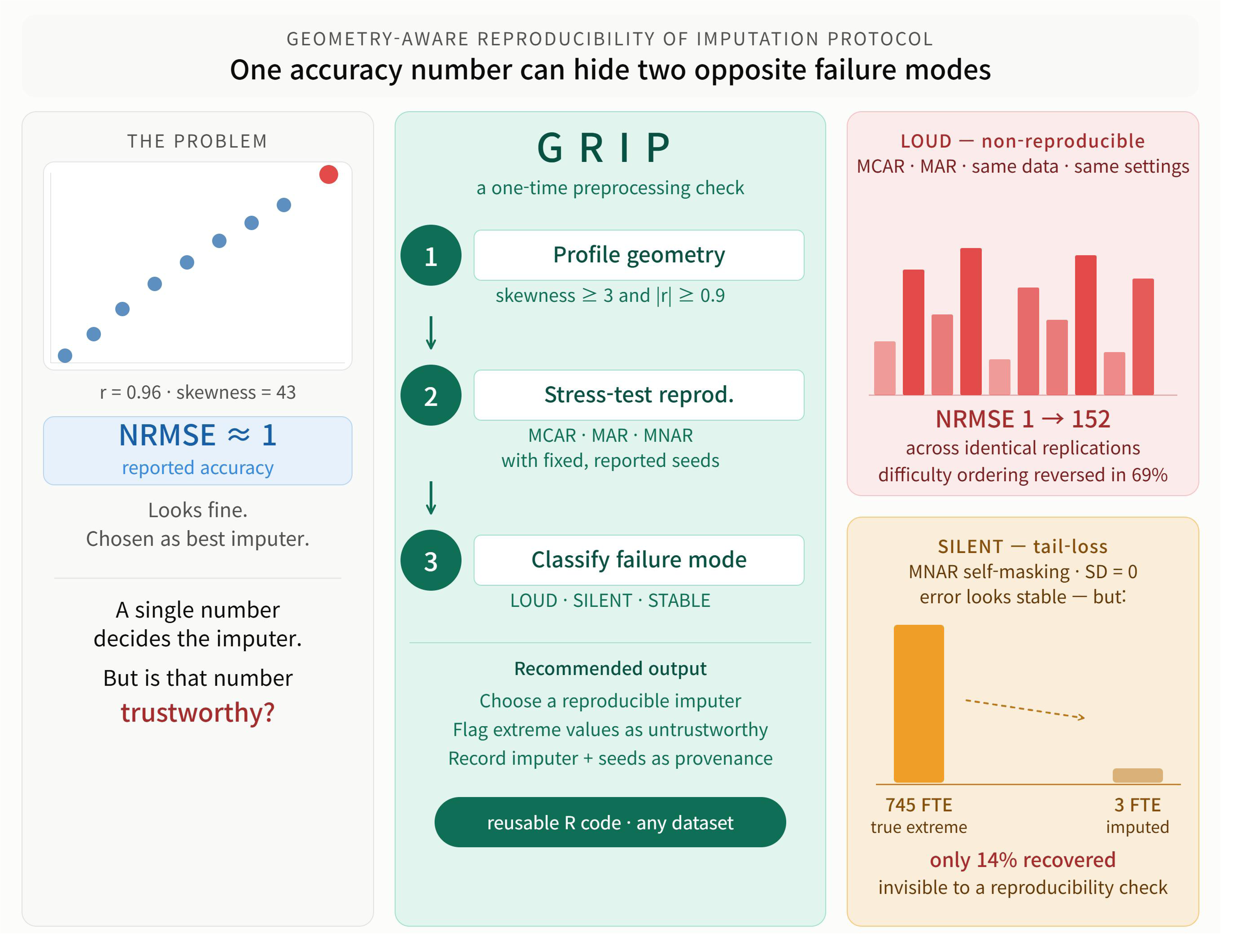

## Background

Imputation replaces missing entries with estimated values and is among the most common preprocessing steps applied to electronic health record (EHR), registry, and administrative datasets before analysis or model training [1, 2]. Imputed values become inputs to every downstream step; the choice of imputer therefore affects the validity and auditability of the results that follow [3]. Missing data is handled inconsistently and poorly in prediction model studies [23, 24], and a consensus standardized evaluation framework is still emerging [10].

In practice, an imputation method is chosen by a single number. An analyst withholds a fraction of observed values, imputes them, and computes an accuracy metric — the normalized root mean squared error (NRMSE) for numeric variables or the proportion of falsely classified entries (PFC) for categorical variables — then selects the method with the lowest value [2, 5]. This workflow treats the accuracy figure as a fixed property. In fact, it is one draw from a distribution induced by which cells happen to be missing. If that distribution is wide, the ranking built on it may not hold on the next draw.

This concern is most acute for heavy-tailed, near-collinear counts: laboratory values, utilization tallies, costs, and staffing. For exactly this geometric profile, the conventional single-accuracy workflow can mislead in two opposite ways. In the first, repeated random deletion of the same variable yields sharply different error values — the reported number is essentially arbitrary. In the second, the accuracy figure is stable, but the variable’s extreme values have been systematically erased. A reproducibility check alone cannot detect the second failure; a magnitude check alone is fooled by both.

This paper introduces GRIP, the Geometry-aware Reproducibility of Imputation Protocol. GRIP (i) profiles each variable’s distributional geometry and collinearity to flag at-risk variables prospectively, (ii) stress-tests reproducibility across MCAR, MAR, and MNAR mechanisms with fixed seeds, and (iii) classifies the failure mode so that an appropriate imputer and an explicit caveat on extreme values can be chosen. Three contributions follow: GRIP itself as a reusable preprocessing protocol; identification and mechanistic explanation of two distinct failure modes unified by one condition; and a synthetic grid establishing that the failure-mode boundary is imputer-class-specific.

## Methods

### The GRIP protocol

GRIP is a three-step, method-agnostic diagnostic intended to run once before an imputer is fixed for downstream analysis (Fig. 1; Additional file 1; Algorithm 1).

**Figure 1.**
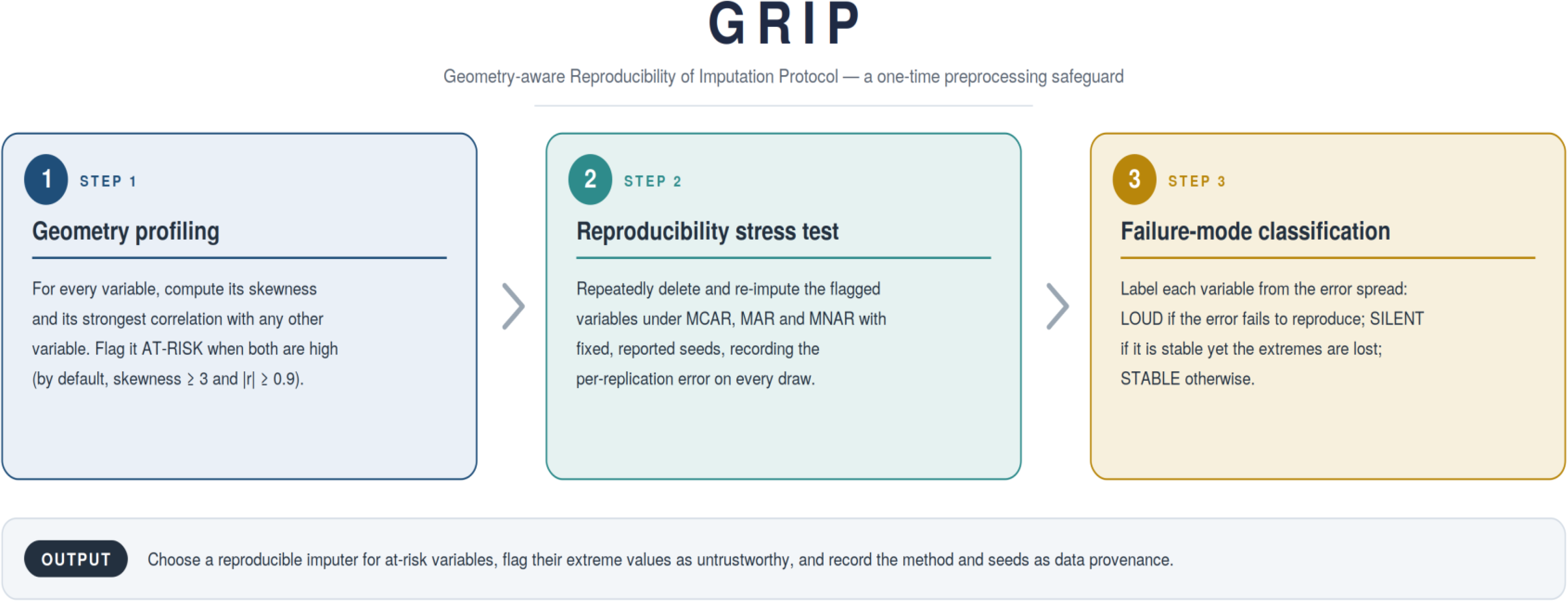
The GRIP protocol. Step 1 profiles each variable’s skewness and collinearity to flag at-risk variables; Step 2 stress-tests reproducibility across MCAR, MAR, and MNAR with fixed seeds; Step 3 classifies each variable’s failure mode (LOUD, SILENT, or STABLE) and yields an actionable preprocessing decision.

**Step 1 — Geometry profiling.** For every variable, GRIP computes sample skewness (tail heaviness) and the maximum absolute Pearson correlation with any other variable (collinearity). A variable is flagged *at-risk* when both exceed user-set thresholds (default: skewness ≥ 3 and |r| ≥ 0.9). The |r| threshold specifically targets the LOUD failure mode, for which a high collinear partner correlation is required to create the instability gamble under MCAR. SILENT failure under MNAR can occur at high skewness regardless of |r| for non-extrapolating imputers; the stress test in Step 2 detects it for any variable.

**Step 2 — Reproducibility stress test.** GRIP repeats a delete-and-impute cycle many times with fixed, reported random seeds under MCAR (uniform random deletion), MAR (deletion probability increasing in an adjacent variable’s rank), and MNAR (self-masking: deletion probability increasing in the variable’s own rank). Each replication records per-variable NRMSE against the withheld ground truth. Testing all three mechanisms is essential: the failure mode differs qualitatively across them, so any single mechanism gives an incomplete picture.

**Step 3 — Failure-mode classification.** For each variable × method × mechanism, GRIP summarizes the NRMSE distribution and assigns one of three labels. *Loud (non-reproducible)*: high across-replication SD. *Silent (tail-loss)*: low SD but extreme values systematically deleted and unrecovered, detectable because the at-risk geometry flagged in Step 1 implies that stable near-floor error has been achieved only by not attempting the extremes. *Stable*: low SD with error at the expected floor. Output: choose a reproducible imputer, mark extreme values as untrustworthy, record the imputer parameters and seeds as data provenance.

The simulation study design follows the ADEMP framework (Aims, Data-generating mechanisms, Estimands, Methods, Performance measures) [22]. A completed ADEMP checklist is provided as Additional file 2.

### Demonstration dataset

GRIP was demonstrated on United States CMS home-health data for calendar year 2017, assembled to operationalize a theory of optimal safe nurse staffing [11–13]. Nursing-quality data were obtained from CMS Home Health Compare; staffing and cost data were obtained from home health agency cost reports and the utilization and payment public-use files, matched and merged by CMS Certification Number during 2021–2022 and analyzed during 2023–2025. From the 2017 file (8,529 agencies, 440 variables), 17 theory-derived variables were selected; retaining cases complete on all yielded 1,885 agencies as ground truth (15 numeric variables, one categorical).

### Imputation methods evaluated

Four imputers were compared: missForest (random-forest single imputation; 100 trees) [5] as the representative high-accuracy single imputer; votingForest (m = 5 random-forest draws collapsed by modal vote) [3] as an internally aggregating single imputer; and mean and median imputation as central-tendency baselines. These four methods separate single-run accuracy from across-replication reproducibility.

### Experimental design and metrics

The delete-and-impute cycle was run under MCAR, MAR, and MNAR at missingness rates of 10%, 20%, and 30% with fixed seeds (MCAR and MAR: 30 replications per rate; MNAR: 10 replications). NRMSE was computed as the square root of mean squared error divided by the variance of the true values; lower values indicate greater accuracy. The across-replication sample SD was the index of irreproducibility. Two supplementary metrics — mean absolute error (MAE) and interquartile-range (IQR)-normalized NRMSE — address the concern that NRMSE instability reflects the variance denominator rather than genuine imputation failure (Additional file 3, Section 10). Within any one variable, the denominator is fixed across replications, so NRMSE variability is driven entirely by the numerator. For right-skewed variables, IQR < SD, so IQR-normalized NRMSE amplifies rather than attenuates the instability signal.

### Synthetic grid

A supplementary k-NN (k = 3) grid was run across 20 combinations of skewness (σ ∈ {0.3, 0.7, 1.1, 1.5}; sample skewness 0.8–13) and Pearson correlation (ρ ∈ {0.50, 0.70, 0.85, 0.95, 0.99}), with n = 500 cases, 30 MCAR and 10 MNAR replications at 20% missingness. The k-NN imputer was chosen as a proxy for non-extrapolating tree-based imputers: it cannot predict beyond the range of its training values. Additional file 4 provides an equivalent grid using missForest. The OLS-based grid characterizing extrapolating-imputer behaviour is presented in Table 4.

### Ethics approval

This study used publicly available, de-identified secondary data and was approved by the Institutional Review Board of the Korea National Institute for Bioethics Policy (approval number P01-202101-22-005; approved 29 January 2021, valid for 10 years). All code is released under the MIT license.

## Results

### Geometry profiling prospectively flags two at-risk variables

Geometry profiling (Table 1) identified exactly two variables meeting both at-risk criteria: Direct Nursing Service Total FTE (skewness 38.6, max |r| 0.96) and Home Health Aide Total FTE (skewness 43.3, max |r| 0.96), which are collinear with each other (r = 0.96). Neither heavy skew alone nor high collinearity alone was sufficient. Total Labor Costs (HH Aide) is comparably skewed (20.8) but weakly collinear (0.28); patient-count variables are moderately collinear (0.84) but only mildly skewed. All three remained reproducible (SD ≤ 0.3). Only the two variables combining extreme skew with near-collinearity showed large SD under random deletion (14.5 and 27.4).

**Table 1.** GRIP Step 1 geometry profile of the 15 numeric variables, with across-replication SD of single-missForest NRMSE from Step 2 (sorted by MCAR SD)

| Variable | Skewness | Max | r |  | SD (MCAR) |
| --- | --- | --- | --- | --- | --- |
| Home Health Aide Total FTE | 43.3 | 0.96 | 27.35 | 30.02 | **Yes** |
| Direct Nursing Service Total FTE | 38.6 | 0.96 | 14.46 | 0.00 | **Yes** |
| Total Labor Costs (HH Aide) | 20.8 | 0.28 | 0.27 | 0.02 | No |
| No. Patients (HH Aide) | 8.0 | 0.84 | 0.13 | 0.16 | No |
| Total Labor Costs (Skilled Nursing) | 8.4 | 0.69 | 0.12 | 0.07 | No |
| Asset Size, Movable Equipment | 8.4 | 0.58 | 0.09 | 0.10 | No |
| Asset Size, Buildings & Fixtures | 9.6 | 0.60 | 0.09 | 0.03 | No |
| No. Patients (Skilled Nursing) | 6.3 | 0.84 | 0.09 | 0.06 | No |
| Admitted to Hospital | 0.6 | 0.31 | 0.03 | 0.02 | No |
| HHCAHPS Summary Star Rating | −0.4 | 0.23 | 0.03 | 0.02 | No |
| Medicare Spending vs National | 0.1 | 0.31 | 0.03 | 0.03 | No |
| Remained in Community after HH | −0.8 | 0.25 | 0.03 | 0.03 | No |
| Quality of Patient Care Star Rating | −0.1 | 0.25 | 0.03 | 0.01 | No |
| Average HCC Score | 0.6 | 0.23 | 0.03 | 0.03 | No |
| Unplanned ER Care | 0.4 | 0.10 | 0.02 | 0.02 | No |

*Note.* Max $|r|$ = maximum absolute Pearson correlation with any other variable. SD = across-replication sample SD ( $n-1$ ) of single-missForest NRMSE over 30 replications × 3 rates. At-risk threshold: skewness $\geq 3$ and max $|r| \geq 0.9$ . FTE = full-time equivalent; HH = home health; ER = emergency room; HCC = hierarchical condition category.

The flag is not an artifact of the chosen thresholds. The two FTE counts have collinearity 0.964; every other variable has maximum |r| ≤ 0.838. Any skewness threshold from 2.5 to 10 combined with any collinearity threshold from 0.85 to 0.95 yields the identical at-risk set. The bimodal SD gap (≤ 0.27 versus ≥ 14.5) cannot be eliminated by removing 19 agencies (top 1%); Additional file 3, Section 8 provides the formal outlier sensitivity analysis.

### Loud non-reproducibility under MCAR and MAR

For the two at-risk variables, missForest accuracy was not reproducible (Table 2; Fig. 2a). Under MCAR, NRMSE for Home Health Aide FTE ranged from approximately 1 to 152 across otherwise identical replications (SD 27.4; median 10.9); for Direct Nursing FTE, from below 1 to 66 (SD 14.5). The instability reversed the difficulty ordering of the two counts in 69% of MCAR replications — whether missForest ranked one variable above the other was determined by the random missingness draw.

**Figure 2.**
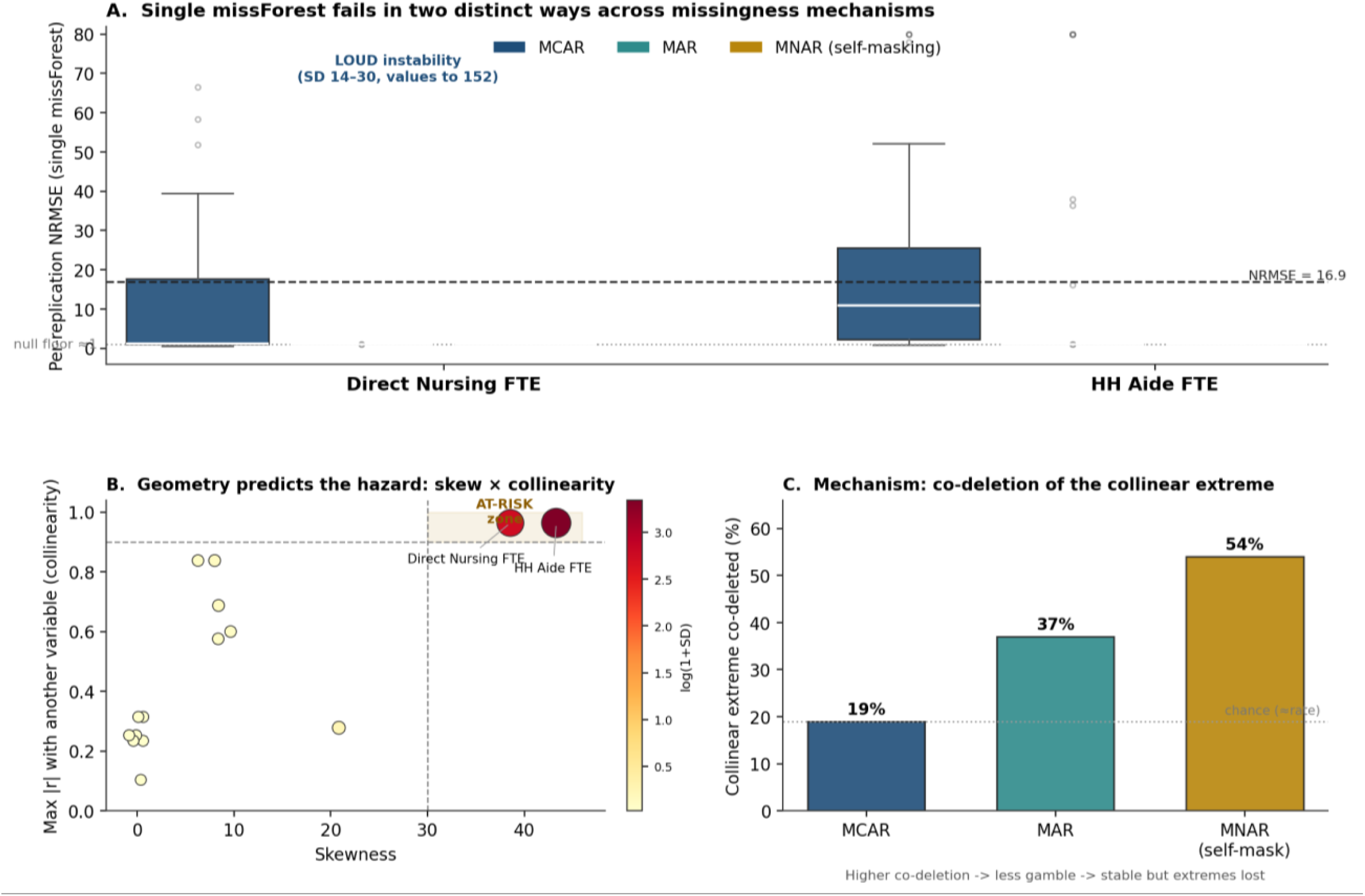
GRIP diagnosis on CMS Home Health calendar year 2017 data (n = 1,885). (A) Per-replication NRMSE for single missForest on the two at-risk FTE counts under MCAR, MAR, and MNAR: LOUD instability under MCAR and MAR (values to 152) versus silent collapse to the null floor under MNAR. (B) Geometry profile: only variables combining high skewness and high collinearity (the at-risk zone) destabilize. All other variables remain in the stable region. (C) Co-deletion of the collinear extreme rises from chance (approximately 19%) under MCAR to approximately 54% under MNAR self-masking (exact binomial test versus chance, p < 10^−15), extinguishing the instability but simultaneously removing the extremes.

**Table 2.** GRIP Step 2 distribution of per-replication NRMSE for the two at-risk variables, by missingness mechanism and imputation method (median; across-replication SD; maximum)

| Mechanism | Method | Dir.<br>median | Dir.<br>SD | Dir.<br>max | HH<br>median | HH<br>SD | HH<br>max |
| --- | --- | --- | --- | --- | --- | --- | --- |
| MCAR | missForest | 1.14 | **14.46** | 66.5 | 10.90 | **27.35** | 152.4 |
|  | votingForest | 1.00 | 5.19 | 50.1 | 1.00 | 0.29 | 3.3 |
|  | mean | 1.01 | 2.26 | 8.4 | 1.89 | 2.08 | 11.0 |
|  | median | 1.00 | 0.02 | 1.1 | 1.01 | 0.01 | 1.1 |
| MAR | missForest | 1.00 | 0.00 | 1.0 | 1.00 | **30.02** | 152.4 |
|  | votingForest | 1.00 | 0.00 | 1.0 | 1.00 | **19.63** | 140.4 |
|  | mean | 1.00 | 0.00 | 1.0 | 1.00 | 0.09 | 1.4 |
|  | median | 1.00 | 0.00 | 1.0 | 1.00 | 0.02 | 1.1 |
| MNAR | missForest | 1.00 | 0.00 | 1.0 | 1.00 | 0.00 | 1.0 |
|  | votingForest | 1.00 | 0.00 | 1.0 | 1.00 | 0.00 | 1.0 |
|  | mean | 1.00 | 0.00 | 1.0 | 1.00 | 0.00 | 1.0 |
|  | median | 1.00 | 0.00 | 1.0 | 1.00 | 0.00 | 1.0 |

*Note.* Dir. = Direct Nursing Service Total FTE; HH = Home Health Aide Total FTE. SD = across-replication sample SD (n-1); max = maximum per-replication NRMSE. Pooled over missingness rates 10/20/30%. Bold SD marks non-reproducible (LOUD) cells (SD > 2). NRMSE ≈ 1 indicates error at the null floor of central-value imputation. VotingForest m = 5.

Under MAR (deletion driven by the collinear partner), instability migrated to whichever count had its extremes deleted while the partner remained informative. Home Health Aide FTE retained SD 30.0; Direct Nursing FTE collapsed to the floor (SD 0.00). In both mechanisms, the loud failure was confined to the at-risk pair; no other variable showed appreciable instability. The bimodal SD gap between at-risk and stable variables is preserved under alternative metrics: the denominator var[x_true] is fixed across replications within each variable, so NRMSE variability is driven entirely by imputation errors.

### Silent tail-loss under MNAR

Under MNAR self-masking, instability vanished: SD fell to 0.00 for both at-risk variables and every method, with NRMSE pinned at the floor. A reproducibility check alone would classify this case as unproblematic.

Tail recovery measured on deleted cells in the upper decile of each variable revealed severe mis-imputation. MissForest recovered on average only 14% of true extreme magnitudes; 99% of extreme values were imputed below half their true value; the mean true extreme of roughly 745 home-health aide FTE was imputed as approximately 3. Stability was achieved not by recovering extreme values but by uniformly regressing them to the centre — a systematic, silent downward bias in exactly the high-volume agencies where understaffing translates most directly into preventable patient harm.

### Unifying mechanism: co-deletion

The two failure modes share one mechanism (Fig. 2c). Instability arises only when an extreme value is missing while its collinear partner still carries the signal. Under MCAR, co-deletion (the proportion of deleted extreme cells for which the collinear partner was also deleted) occurred at the chance rate (approximately 19%; exact binomial test, p = 0.71). The partner remained informative and the gamble was live — LOUD instability. Under MNAR self-masking, co-deletion rose to approximately 54% (2.7 times the chance rate; p < 10⁻¹⁵), because self-masking selects the same extreme agencies in both collinear columns simultaneously. The gamble was extinguished, but so was tail recoverability — SILENT tail-loss. MAR occupied the intermediate regime (co-deletion approximately 37%).

### Conventional evaluation and stable imputers

An independent five-replication out-of-bag run (Table 3) illustrates the practical consequence. Direct Nursing FTE NRMSE spanned 0.67–10.9 within five replications, with the larger error alternating between the two variables across replications. A bootstrap of five-replication means assigns roughly 15% probability to a mean of 16.9 or higher, while only approximately 28% of individual replications reach 16.9 at all.

**Table 3.** An independent five-replication out-of-bag run for the two at-risk variables, illustrating that a conventional five-replication evaluation is non-reproducible.

| Replication | Direct Nursing FTE NRMSE | HH Aide FTE NRMSE |
| --- | --- | --- |
| 1 | 7.66 | 0.99 |
| 2 | 9.90 | 4.23 |
| 3 | 10.88 | 4.97 |
| 4 | 8.22 | 1.00 |
| 5 | 0.67 | 28.99 |
| **Mean (SD)** | **7.5 (4.0)** | **8.0 (11.9)** |

*Note.* Out-of-bag NRMSE from single missForest under MCAR-style random deletion at 20% rate. Within five replications, the Direct value spans 0.67–10.9 and the HH value spans 0.99–29.0, with the larger error alternating between the two variables across replications.

**Table 4.**
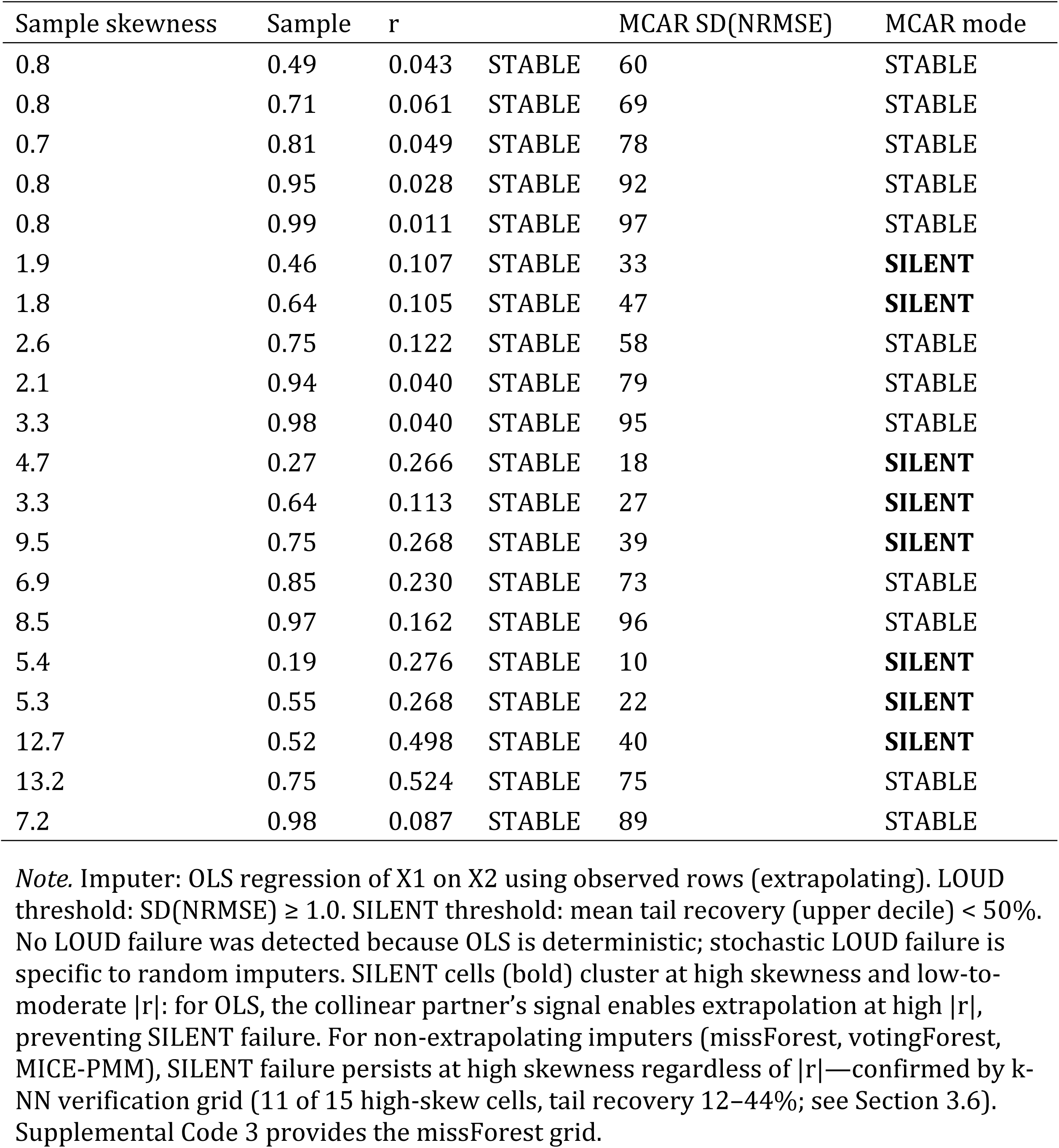
Synthetic grid results: failure-mode classification across 20 (skewness, |r|) combinations, with OLS imputation (n = 500; 20% missingness; 30 MCAR, 10 MNAR replications)

| Sample skewness | Sample | r |  | MCAR SD(NRMSE) | MCAR mode |
| --- | --- | --- | --- | --- | --- |
| 0.8 | 0.49 | 0.043 | STABLE | 60 | STABLE |
| 0.8 | 0.71 | 0.061 | STABLE | 69 | STABLE |
| 0.7 | 0.81 | 0.049 | STABLE | 78 | STABLE |
| 0.8 | 0.95 | 0.028 | STABLE | 92 | STABLE |
| 0.8 | 0.99 | 0.011 | STABLE | 97 | STABLE |
| 1.9 | 0.46 | 0.107 | STABLE | 33 | **SILENT** |
| 1.8 | 0.64 | 0.105 | STABLE | 47 | **SILENT** |
| 2.6 | 0.75 | 0.122 | STABLE | 58 | STABLE |
| 2.1 | 0.94 | 0.040 | STABLE | 79 | STABLE |
| 3.3 | 0.98 | 0.040 | STABLE | 95 | STABLE |
| 4.7 | 0.27 | 0.266 | STABLE | 18 | **SILENT** |
| 3.3 | 0.64 | 0.113 | STABLE | 27 | **SILENT** |
| 9.5 | 0.75 | 0.268 | STABLE | 39 | **SILENT** |
| 6.9 | 0.85 | 0.230 | STABLE | 73 | STABLE |
| 8.5 | 0.97 | 0.162 | STABLE | 96 | STABLE |
| 5.4 | 0.19 | 0.276 | STABLE | 10 | **SILENT** |
| 5.3 | 0.55 | 0.268 | STABLE | 22 | **SILENT** |
| 12.7 | 0.52 | 0.498 | STABLE | 40 | **SILENT** |
| 13.2 | 0.75 | 0.524 | STABLE | 75 | STABLE |
| 7.2 | 0.98 | 0.087 | STABLE | 89 | STABLE |

*Note.* Imputer: OLS regression of X1 on X2 using observed rows (extrapolating). LOUD threshold: SD(NRMSE) ≥ 1.0. SILENT threshold: mean tail recovery (upper decile) $<$ 50%. No LOUD failure was detected because OLS is deterministic; stochastic LOUD failure is specific to random imputers. SILENT cells (bold) cluster at high skewness and low-to-moderate |r|: for OLS, the collinear partner's signal enables extrapolation at high |r|, preventing SILENT failure. For non-extrapolating imputers (missForest, votingForest, MICE-PMM), SILENT failure persists at high skewness regardless of |r|—confirmed by k-NN verification grid (11 of 15 high-skew cells, tail recovery 12–44%; see Section 3.6). Supplemental Code 3 provides the missForest grid.

Within the same stress test, votingForest and median imputation were reproducible for the at-risk variables under MCAR (SD ≤ 0.3). VotingForest’s per-cell modal vote supplies the aggregation that single missForest lacks; any central-tendency or aggregating rule restores stability. NRMSE ≈ 1 signifies only that stable methods do not destabilize while declining to attempt the extremes. Under MAR, even votingForest inherited residual instability on Home Health Aide FTE (SD 19.6), indicating that no single imputer fully escapes the problem — reinforcing the case for a diagnostic safeguard rather than reliance on a fixed method.

### Synthetic grid: failure-mode boundary is imputer-class-specific

The OLS grid (Table 4) established the baseline for extrapolating imputers. Under MNAR, SILENT failure occurred in 8 of 20 cells — all at high skewness combined with |r| < 0.85. At high correlation, OLS extrapolated using the collinear partner’s signal to recover extremes (STABLE). Under MCAR, no LOUD failure was detected (max SD = 0.53); OLS is deterministic, and LOUD failure requires stochastic imputer behaviour.

The k-NN verification grid confirmed that for non-extrapolating imputers, the pattern inverts. SILENT failure under MNAR occurred in 11 of 15 high-skew cells (skewness > 3), with tail recovery ranging from 12% to 44%, regardless of correlation level. The four borderline cells all had |r| ≥ 0.97; at near-unity correlation, k-NN can locate the extreme training point as an exact nearest neighbour — a precision not available to random-forest trees. MCAR instability was elevated for high-skew combinations (SD up to 0.39) compared with low-skew stable combinations (SD ≤ 0.11); reaching the SD ≥ 1.0 LOUD threshold requires both the extreme skewness of the CMS data (38–43) and the specific stochasticity of the random-forest algorithm.

## Discussion

For heavy-tailed, collinear variables, one imputation-accuracy number is a draw from a distribution whose shape depends on data geometry and the missingness mechanism. In this demonstration it failed in two opposite ways: non-reproducible under MCAR and MAR (LOUD), stably misleading under MNAR (SILENT). The two modes are unified by one condition — instability requires an extreme value to be absent while its collinear partner remains observed — and are toggled by the missingness mechanism via the co-deletion rate. GRIP operationalizes this insight as a reusable preprocessing protocol.

These findings refine, rather than contradict, asymptotic guidance that mean-style imputation is optimal for downstream prediction [15]. Asymptotic optimality concerns expectations; it is silent about the reproducibility of a single estimate under finite, heavy-tailed, collinear data. Where asymptotics are least informative, GRIP is most needed. The findings also complement reports that the amount of missingness can matter more than the mechanism for predictive performance [17]: for the reproducibility of the evaluation itself, the mechanism is decisive — it sets the co-deletion rate that toggles between failure modes. Missing data is handled poorly and inconsistently in prediction model studies [23, 24]; GRIP’s auditable record addresses this gap directly.

A reproducibility check without geometry profiling would find SD ≈ 0 under MNAR and classify the at-risk variables as safe — missing the silent tail-loss entirely. Geometry profiling without a stress test would flag the variables but could not reveal how they fail or which imputer to trust. The combination of prospective geometric flagging, multi-mechanism stress testing, and explicit failure-mode classification is required to detect both modes. This is why GRIP is framed as a protocol rather than a single statistic.

The synthetic grid establishes that the failure-mode boundary is imputer-class-specific. For OLS (extrapolating), SILENT failure under MNAR is concentrated at high skewness and low correlation, where extrapolation power depends on the collinear signal. For non-extrapolating imputers — all tree-based methods including missForest, votingForest, and MICE with predictive mean matching (MICE-PMM) — SILENT failure occurs at high skewness regardless of correlation, because the imputer cannot predict beyond its training range when all extreme training cases are deleted under self-masking. The k-NN grid confirmed this pattern empirically (11 of 15 high-skew combinations; tail recovery 12–44%). The Step 1 |r| threshold targets the LOUD failure zone and the zone of most severe MNAR co-deletion; for SILENT prevention with non-extrapolating imputers, high skewness alone is the primary indicator, and the stress test in Step 2 is the definitive arbiter.

The at-risk geometry — extreme right-skew with near-collinearity — is common across biomedical informatics: laboratory results, utilization tallies, billing amounts, length-of-stay, and device-derived signals all share this profile. The same two failure modes are hypothesized wherever this geometry occurs, for imputers of the respective extrapolation class. This is a hypothesis supported by one dataset with two at-risk variables; confirming its prevalence requires multi-dataset evaluation, for which Additional file 4 provides a replicable framework.

The implications for high-stakes data pipelines are immediate. When workforce policy is built on a non-reproducible accuracy figure, the uncertainty of that figure is inherited by every staffing target derived from it. GRIP yields an auditable record — flagged variables, mechanism-specific reproducibility, chosen imputer, seeds — that supports reproducible, accountable pipelines. The same logic extends to any variable carrying irreducible, decision-bearing value in its extremes: peak blood pressure, maximum administered dose, fall counts, or longest length of stay.

### Limitations

The evidence is a demonstration, not a validation: the failure modes are established on one dataset with two at-risk variables. The mechanism is analytically explained and reproduced on a synthetic grid, which distinguishes the finding from a single-dataset anecdote. Only one MNAR process was examined; the silent mode should be read as one demonstrated possibility under MNAR. Skewness is sensitive to the largest observations; a formal outlier sensitivity analysis is provided in Additional file 3, Section 8. The default thresholds are practical starting values; the stress test, not the flag, is the arbiter. VotingForest has not been independently peer-reviewed; no conclusions depend on it. GRIP diagnoses reproducibility and tail-recoverability but does not by itself repair tail-loss. The demonstration data are agency-level administrative records; confirming the same behaviour in patient-level EHR data is part of the validation called for here.

## Conclusions

Imputation is typically chosen by one accuracy number. For the heavy-tailed, collinear variables that pervade biomedical data, that number can mislead in two opposite ways: non-reproducible (LOUD) or stably misleading while extreme values are silently lost (SILENT). GRIP surfaces both, by profiling data geometry, stress-testing reproducibility across MCAR, MAR, and MNAR, and classifying the failure mode. Applied to real home-health staffing data, GRIP prospectively isolated the two at-risk variables, showed that a conventional few-replication accuracy figure is one draw from a wide, unstable distribution, and identified the silent MNAR bias that a reproducibility check alone would miss. A k-NN verification grid confirmed that SILENT failure under MNAR is present at high skewness regardless of correlation for non-extrapolating imputers. GRIP is available as open R code and formal pseudocode (Additional file 1) and can be run on any dataset immediately, using only the data, a small set of candidate imputers, and the provided code.

## Declarations

### Ethics approval and consent to participate

This study used publicly available, de-identified secondary data and was approved by the Institutional Review Board of the Korea National Institute for Bioethics Policy (approval number P01-202101-22-005; approved 29 January 2021, valid for 10 years). As the data are de-identified and publicly available, individual consent to participate was not required.

### Consent for publication

Not applicable.

### Availability of data and materials

The Centers for Medicare & Medicaid Services (CMS) Home Health datasets analysed in this study are publicly available from CMS (https://www.cms.gov). The derived analysis-ready dataset (1,885 complete cases) is available from the corresponding author upon reasonable request. The GRIP implementation is provided as Additional file 1 (project name: GRIP; programming language: R; license: MIT; no restrictions on use by non-academics). All experiment code, including the synthetic grid, is provided as Additional files 3–4 under the MIT license.

### Authors’ contributions

CSYP conceived the study, developed the GRIP methodology, designed the screening protocol, curated and analyzed the data, developed the software, validated the findings, prepared the visualizations, and wrote and revised the manuscript. The author read and approved the final manuscript.

## Supporting information

Additional File_Algorithm 1

Additional File 1_Code_1_GRIP

Additional File 2_ADEMP

Additional File 3_Code_2_Experiments

Additional File 4_Code_3_Grid

## Data Availability

The Centers for Medicare & Medicaid Services (CMS) Home Health datasets analysed in this study are publicly available from CMS (https://www.cms.gov). The derived analysis-ready dataset (1,885 complete cases) is available from the corresponding author upon reasonable request. The GRIP implementation is provided as Additional file 1 (project name: GRIP; programming language: R; license: MIT; no restrictions on use by non-academics). All experiment code, including the synthetic grid, is provided as Additional files 3-4 under the MIT license.

## Acknowledgements

The author thanks Hyoungchul Choi, MS (LG CNS Co., Ltd., Seoul, Republic of Korea), the original developer of votingForest, for sharing the original implementation and for helpful discussion.

## Use of generative AI

During the preparation of this work, the author used a generative AI assistant (Claude, Anthropic) solely for language editing and refinement. After using this tool, the author reviewed and edited the content as needed and takes full responsibility for the content of the publication.

## Declarations

### CRediT author statement

Claire Su-Yeon Park: Conceptualization, Methodology, Software, Formal analysis, Investigation, Data curation, Writing – original draft, Writing – review & editing, Visualization. As sole author, she is responsible for all aspects of the work.

### Generative AI disclosure

During the preparation of this work, the author used a generative AI assistant (Claude, Anthropic) solely for language editing and refinement. The author reviewed and verified all content and code and takes full responsibility for the work.

