## Additional File_Algorithm 1 for "Geometry-Aware Reproducibility of Imputation Protocol (GRIP): diagnosing two failure modes of single imputation for heavy-tailed, collinear variables in biomedical data": Additional File_Algorithm 1.docx

### Additional file 1: Algorithm 1 — The GRIP protocol (pseudocode)

**Input:** data D; thresholds θs = 3, θr = 0.9; rates Rk; replications N; seed s0; imputers F = {f1, …, fp}; mechanisms M = {MCAR, MAR, MNAR}

**Output:** per-variable failure-mode label and recommended imputer

**Step 1: Geometry profiling**

```
for each variable v in D:
    skew[v] <- sample_skewness(v)
    coll[v] <- max_{u ≠ v} | corr(v, u) |
    atRisk[v] <- (skew[v] >= θs) AND (coll[v] >= θr)
```

**Step 2: Multi-mechanism reproducibility stress test**

```
CC <- complete_cases(D)
for mech in M, rate in Rk, r in 1..N, v in target_vars:
    seed  <- s0 + r
    set_seed(seed)
    mask  <- amputate(CC, v, rate, mech)    # MCAR / MAR / MNAR rule
    Dmiss <- CC with CC[mask, v] set to NA
    for f in F:
        imp <- f(Dmiss)
        E[mech, rate, seed, v, f] <- NRMSE(imp[v], CC[v], mask)
```

**Step 3: Failure-mode classification**

```
for each (v, f, mech):
    sd_e <- SD_r( E[mech, *, *, v, f] )
    if sd_e >= 1:
        label <- LOUD (non-reproducible)
    elif atRisk[v] AND tail_deleted(mech, v):
        label <- SILENT (tail-loss)
    else:
        label <- STABLE
return labels, recommended reproducible imputer for at-risk v
```

**Notes.** NRMSE = normalized root mean squared error = √(mean[(x_imp − x_true)²] / var[x_true]). The LOUD threshold (SD ≥ 1) was chosen because the observed SD distribution is bimodal—13 variables at or below 0.27 and two at or above 14—so any threshold between these clusters yields the same classification. Default thresholds θs = 3 and θr = 0.9 represent a stable region, not a knife-edge boundary; any skewness threshold from 2.5 to 10 combined with any collinearity threshold from 0.85 to 0.95 yields the identical at-risk set on the CMS demonstration data. The stress test, not the flag, is the arbiter of instability.
