## Additional File 1_Code_1_GRIP for "Geometry-Aware Reproducibility of Imputation Protocol (GRIP): diagnosing two failure modes of single imputation for heavy-tailed, collinear variables in biomedical data": Additional File 1_Code_1_GRIP.R

```
########################################################################
##  GRIP: Geometry-aware Reproducibility of Imputation Protocol
##  A diagnostic procedure to detect non-reproducible and silently
##  biased single imputation for heavy-tailed, collinear variables.
##
##  Reference: Park CS-Y. Geometry-aware Reproducibility of Imputation
##  Protocol (GRIP): diagnosing two failure modes of single imputation
##  for heavy-tailed, collinear variables in biomedical data. (2026)
##
##  Author: Claire Su-Yeon Park (ORCID 0000-0002-2109-9885)
##  License: MIT.  Dependencies: stats (base), optional: missForest, e1071
########################################################################

## ----------------------------------------------------------------------
## STEP 1 — GEOMETRY PROFILING
## Flag variables whose geometry (heavy right-skew AND high collinearity
## with another variable) places them in the at-risk zone.
## ----------------------------------------------------------------------
grip_profile <- function(data, skew_thr = 3, collinearity_thr = 0.9) {
  num <- data[, sapply(data, is.numeric), drop = FALSE]
  sk  <- apply(num, 2, function(x) {
            x <- x[!is.na(x)]; m <- mean(x)
            mean((x - m)^3) / (mean((x - m)^2)^1.5)   # sample skewness
         })
  R   <- abs(stats::cor(num, use = "pairwise.complete.obs"))
  diag(R) <- 0
  maxr <- apply(R, 2, max, na.rm = TRUE)
  at_risk <- (sk >= skew_thr) & (maxr >= collinearity_thr)
  data.frame(variable = names(sk), skewness = round(sk, 2),
             max_abs_r = round(maxr, 3), at_risk = at_risk,
             row.names = NULL)
}

## ----------------------------------------------------------------------
## STEP 2 — MULTI-MECHANISM REPRODUCIBILITY STRESS TEST
## Repeatedly delete-and-impute under MCAR, MAR, and MNAR with fixed,
## reported seeds; record per-replication NRMSE for each method/variable.
## `impute_fns` is a named list of functions f(data_with_NA) -> completed.
## ----------------------------------------------------------------------
.amputate <- function(X, j, rate, mechanism, partner = NULL, seed) {
  set.seed(seed); n <- nrow(X); k <- round(rate * n); M <- logical(n)
  if (mechanism == "MCAR") {
    M[sample.int(n, k)] <- TRUE
  } else if (mechanism == "MAR") {                 # depends on PARTNER value
    s <- rank(X[, partner]) + rnorm(n, 0, n * 0.05)
    M[order(s, decreasing = TRUE)[seq_len(k)]] <- TRUE
  } else if (mechanism == "MNAR") {                # self-masking (own value)
    s <- rank(X[, j]) + rnorm(n, 0, n * 0.05)
    M[order(s, decreasing = TRUE)[seq_len(k)]] <- TRUE
  }
  M
}

nrmse <- function(imp, truth, mask)
  sqrt(mean((imp[mask] - truth[mask])^2) / stats::var(truth[mask]))

## Tail-recoverability on the deleted extreme cells (true value in the upper
## `q` quantile): returns the mean imputed/true magnitude ratio and the
## fraction imputed below half the true value.  Low recovery + low across-
## replication SD is the signature of the SILENT (tail-loss) failure mode.
tail_recovery <- function(imp, truth, mask, q = 0.90) {
  thr <- stats::quantile(truth, q, na.rm = TRUE)
  idx <- which(mask & truth >= thr)
  if (!length(idx)) return(c(recovered = NA, frac_below_half = NA))
  c(recovered       = mean(imp[idx] / truth[idx]),
    frac_below_half = mean(imp[idx] < 0.5 * truth[idx]))
}

grip_stress <- function(data, target_vars, impute_fns,
                        mechanisms = c("MCAR", "MAR", "MNAR"),
                        rates = c(.10, .20, .30), n_rep = 30,
                        seed0 = 1001) {
  cc  <- data[stats::complete.cases(data), , drop = FALSE]
  out <- list(); row <- 1
  for (mech in mechanisms)
   for (rate in rates)
    for (r in seq_len(n_rep)) {
      seed <- seed0 + r
      for (v in target_vars) {
        partner <- names(which.max(abs(stats::cor(cc)[v, ][setdiff(names(cc), v)])))
        M <- .amputate(cc, v, rate, mech, partner, seed)
        D <- cc; D[M, v] <- NA
        for (fn in names(impute_fns)) {
          imp <- impute_fns[[fn]](D)
          out[[row]] <- data.frame(mechanism = mech, rate = rate, seed = seed,
                                   variable = v, method = fn,
                                   nrmse = nrmse(imp[[v]], cc[[v]], M),
                                   prop_codeleted = NA)
          row <- row + 1
        }
      }
    }
  do.call(rbind, out)
}

## ----------------------------------------------------------------------
## STEP 3 — FAILURE-MODE CLASSIFICATION
## For each variable x method x mechanism, classify into:
##   STABLE            : low across-replication SD AND error near null floor
##   LOUD (unstable)   : high across-replication SD  (non-reproducible)
##   SILENT (tail-loss): low SD but systematic extreme-value error
## ----------------------------------------------------------------------
grip_classify <- function(stress, sd_thr = 1, floor_band = c(0.5, 1.5),
                          tail_loss_check = NULL) {
  agg <- aggregate(nrmse ~ mechanism + variable + method, stress,
                   function(x) c(sd = sd(x), med = median(x)))
  agg <- do.call(data.frame, agg)
  names(agg)[4:5] <- c("sd", "median")
  agg$failure_mode <- with(agg, ifelse(
      sd >= sd_thr, "LOUD (non-reproducible)",
      ifelse(!is.null(tail_loss_check) & tail_loss_check,
             "SILENT (tail-loss)", "STABLE")))
  agg[order(-agg$sd), ]
}

## ----------------------------------------------------------------------
## REFERENCE IMPUTERS (illustrative)
## votingForest: m random-forest (missForest-style) draws collapsed PER
## CELL by a modal vote.  TIE / UNANIMITY RULE (per H. Choi):
##   - if all m candidate values are identical, that value is taken;
##   - if a single mode exists, the mode is taken;
##   - if several values tie for the mode, one of the tied modes is taken.
## ----------------------------------------------------------------------
vote_cell <- function(v) {                 # v = m candidate values for one cell
  v <- round(v, 1)
  tab <- table(v)
  modes <- as.numeric(names(tab)[tab == max(tab)])
  if (length(modes) == 1L) return(modes)   # unique mode (covers unanimity)
  modes[order(modes)][ceiling(length(modes) / 2)]   # tie -> central tied mode
}
```
