## Additional File 2_ADEMP for "Geometry-Aware Reproducibility of Imputation Protocol (GRIP): diagnosing two failure modes of single imputation for heavy-tailed, collinear variables in biomedical data": Additional File 2_ADEMP.docx

### Additional file 2: ADEMP reporting checklist

**Reference:** Morris TP, White IR, Crowther MJ. Using simulation studies to evaluate statistical methods. *Statistics in Medicine*. 2019;38(11):2074–2102.

**Study:** Geometry-Aware Reproducibility of Imputation Protocol (GRIP): Diagnosing Two Failure Modes of Single Imputation for Heavy-Tailed, Collinear Variables in Biomedical Data.

The simulation components of this study (synthetic grid, Methods section “Synthetic grid” and Results section “Synthetic grid: failure-mode boundary is imputer-class-specific”) follow the ADEMP framework. Items below are checked (✓) where the reporting requirement is met, with a reference to the relevant manuscript location.

#### A — Aims

| Item | Requirement | Status | Location |
| --- | --- | --- | --- |
| A1 | State the research questions the simulation addresses | ✓ | Introduction para 4; Methods Methods |
| A2 | Clarify whether the study is a Monte Carlo study, benchmark, or comparison | ✓ | Methods: “supplementary k-NN grid … OLS-based grid” — comparison across imputer classes |

#### D — Data-generating mechanism

| Item | Requirement | Status | Location |
| --- | --- | --- | --- |
| D1 | Describe the data-generating mechanism (DGM) fully | ✓ | Methods: Gaussian-copula bivariate lognormal; X1 = exp(σZ1), X2 = exp(σZ2); (Z1,Z2) ~ N(0,[1,ρ;ρ,1]); five N(0,1) nuisance variables |
| D2 | State parameter values and their ranges | ✓ | Methods: σ ∈ {0.3, 0.7, 1.1, 1.5}; ρ ∈ {0.50, 0.70, 0.85, 0.95, 0.99}; n = 500; 20% missingness |
| D3 | State number of replications per scenario | ✓ | Methods: 30 MCAR replications; 10 MNAR replications per cell |
| D4 | State random seeds or seeding strategy | ✓ | Methods: “fixed and reported seeds”; Supplemental Code 2 reports all seeds used |
| D5 | Justify the chosen parameter values with respect to real data | ✓ | Methods: parameter range chosen to span from below-at-risk (σ = 0.3, skew ≈ 0.8) to high-risk (σ = 1.5, skew ≈ 8–21); CMS data (skew 38–43) noted as exceeding grid range |

#### E — Estimands

| Item | Requirement | Status | Location |
| --- | --- | --- | --- |
| E1 | Define clearly what is being estimated | ✓ | Methods / Results: across-replication SD(NRMSE) under MCAR (LOUD index); mean tail-recovery under MNAR (SILENT index) |
| E2 | Distinguish estimands from targets and performance measures | ✓ | Methods: NRMSE defined precisely; SD of NRMSE is the estimand; LOUD/SILENT classification is the decision rule |

#### M — Methods

| Item | Requirement | Status | Location |
| --- | --- | --- | --- |
| M1 | Describe all methods compared (including their implementation) | ✓ | Methods: OLS (extrapolating reference); k-NN k = 3 (non-extrapolating proxy); Supplemental Code 3 for missForest grid |
| M2 | State software and packages used | ✓ | Supplemental Code 1–3: R; Supplemental Code 2 Section 10: Python-equivalent verification. All code released under MIT license |
| M3 | State whether methods are implemented from published software or custom code | ✓ | Methods: “GRIP is implemented as three R functions—grip_profile(), grip_stress(), grip_classify()—provided as Supplemental Code 1” |

#### P — Performance measures

| Item | Requirement | Status | Location |
| --- | --- | --- | --- |
| P1 | Define performance measures and justify their choice | ✓ | Methods: SD(NRMSE) for reproducibility (LOUD); mean tail-recovery ratio for bias toward extremes (SILENT); chosen because they directly map to the two failure modes |
| P2 | State how the performance measures will be estimated | ✓ | Methods: empirical distribution across replications; SD (n−1) as the index of irreproducibility |
| P3 | State LOUD/SILENT classification thresholds and their justification | ✓ | Methods: LOUD if SD ≥ 1.0 (bimodal gap justification in Results); SILENT if tail recovery < 50% |
| P4 | Report uncertainty around performance estimates (e.g., Monte Carlo SE) | Partial | Results reports per-cell results across all 20 combinations; Monte Carlo SEs not separately tabulated but available from Supplemental Code 2 |

#### Additional reporting items

| Item | Requirement | Status | Location |
| --- | --- | --- | --- |
| R1 | Describe real data used to motivate or anchor simulation parameters | ✓ | Methods–2.3: CMS home-health 2017 data; Table 1 reports empirical skewness and correlation values |
| R2 | State any deviations from preregistered protocol | N/A | No preregistration; simulation designed as secondary verification of CMS demonstration |
| R3 | Provide replication code | ✓ | Supplemental Code 2 (Python k-NN grid); Supplemental Code 3 (R missForest grid); released under MIT license |
