## Additional File 3_Code_2_Experiments for "Geometry-Aware Reproducibility of Imputation Protocol (GRIP): diagnosing two failure modes of single imputation for heavy-tailed, collinear variables in biomedical data": Additional File 3_Code_2_Experiments.R

```
########################################################################
##  Supplemental Code 2 (Updated) — Experiment driver
##
##  Park CS-Y. Geometry-Aware Reproducibility of Imputation Protocol
##  (GRIP): Diagnosing Two Failure Modes of Single Imputation for
##  Heavy-Tailed, Collinear Variables in Biomedical Data.
##  Journal of Clinical Epidemiology (submitted).
##
##  Sections:
##    1.  Table 1  — geometry profile
##    2.  Table 2  — NRMSE distribution (missForest, votingForest,
##                   MICE-PMM, mean, median)
##    3.  Sec 4.3  — tail-recovery under MNAR (silent tail-loss mode)
##    4.  Sec 4.4  — co-deletion + exact binomial test
##    5.  Sec 4.1  — threshold-robustness of the at-risk flag
##    6.  Sec 4.5  — bootstrap of the five-replication mean;
##                   difficulty-ordering reversal
##    7.  Table 3  — illustrative five-replication out-of-bag run
##    8.  Sec 4.7  — outlier sensitivity (remove top 1% agencies)  [NEW]
##    9.  Table 2  — supplementary MICE(PMM) rows                  [NEW]
##   10.  Suppl. Table 1 — MAE and IQR-NRMSE alternative metrics   [NEW]
##
##  Author : Claire Su-Yeon Park (ORCID 0000-0002-2109-9885)
##  License: MIT
##
##  Dependencies:
##    install.packages(c("missForest", "e1071", "mice"))
##  Companion:
##    source("Supplemental_Code_1_GRIP.R")
##  Input:
##    rdata_1885.csv  (1,885 complete cases x 15 numeric vars)
##
##  Reproducibility note: all randomness is seeded and seeds are
##  fixed below.  Individual NRMSE values vary across implementations
##  because random-forest imputation is stochastic; the qualitative
##  findings reproduce.
########################################################################

suppressMessages({
  library(missForest)
  library(e1071)
  library(mice)
})
source("Supplemental_Code_1_GRIP.R")

RATES <- c(0.10, 0.20, 0.30)
REPS  <- c(MCAR = 30L, MAR = 30L, MNAR = 10L)
SEED0 <- 1000L
NTREE <- 100L
M_VF  <- 5L    # votingForest draws
M_MI  <- 5L    # MICE imputed datasets (reduced to one by column means)

## ── data ───────────────────────────────────────────────────────────────────
D       <- read.csv("rdata_1885.csv", check.names = FALSE)
num_all <- D[, vapply(D, is.numeric, logical(1)), drop = FALSE]
num     <- num_all[, setdiff(names(num_all), "Type_Ownership_Cost_code"),
                   drop = FALSE]
DIRECT  <- "Direct_Nursing_Service__Total_FTE"
HHAIDE  <- "HH_Aide__Total_FTE"
TARGET  <- c(DIRECT, HHAIDE)
stopifnot(nrow(num) == 1885L, ncol(num) == 15L, all(TARGET %in% names(num)))

## ── impute_one: one column, one method, one masked dataset ─────────────────
impute_one <- function(Dmiss, v, method, seed) {
  truth <- Dmiss[[v]]; M <- is.na(truth)
  if (method == "mean") {
    x <- truth; x[M] <- mean(truth[!M]); return(x)
  }
  if (method == "median") {
    x <- truth; x[M] <- median(truth[!M]); return(x)
  }
  if (method == "missForest") {
    set.seed(seed)
    return(missForest(Dmiss, ntree = NTREE, verbose = FALSE)$ximp[[v]])
  }
  if (method == "votingForest") {
    draws <- vector("list", M_VF)
    for (d in seq_len(M_VF)) {
      set.seed(seed * 100L + d)
      draws[[d]] <- missForest(Dmiss, ntree = NTREE, verbose = FALSE)$ximp[[v]]
    }
    out <- draws[[1]]
    for (i in which(M)) out[i] <- vote_cell(vapply(draws, `[`, numeric(1), i))
    return(out)
  }
  if (method == "mice_pmm") {
    ## M_MI imputed datasets; reduce to one by taking column mean
    set.seed(seed)
    mids <- suppressWarnings(
      mice(Dmiss, m = M_MI, method = "pmm",
           printFlag = FALSE, maxit = 5)
    )
    ## Average imputed values across m datasets for the target column
    imp_mat <- sapply(seq_len(M_MI), function(k)
                 complete(mids, k)[[v]])
    avg_imp <- rowMeans(imp_mat)
    ## Return the full vector (observed + imputed positions)
    x <- truth
    x[M] <- avg_imp[M]
    return(x)
  }
}

## ── alternative metrics ────────────────────────────────────────────────────
mae_metric <- function(imp, truth, mask) {
  mean(abs(imp[mask] - truth[mask]))
}

iqr_nrmse <- function(imp, truth, mask) {
  iqr <- IQR(truth[mask], na.rm = TRUE)
  if (iqr < 1e-10) return(NA_real_)
  sqrt(mean((imp[mask] - truth[mask])^2)) / iqr
}

## ======================================================================
## SECTION 1: TABLE 1 — geometry profile
## ======================================================================
cat("\n=== SECTION 1: TABLE 1: geometry profile ===\n")
prof <- grip_profile(num, skew_thr = 3, collinearity_thr = 0.9)
print(prof[order(-prof$max_abs_r, -prof$skewness), ], row.names = FALSE)

## ======================================================================
## SECTION 2: TABLE 2 — NRMSE (original 4 methods)
## ======================================================================
METHODS_ORIG <- c("missForest", "votingForest", "mean", "median")

rows <- list(); k <- 1L
for (mech in c("MCAR", "MAR", "MNAR")) {
  for (rate in RATES) for (r in seq_len(REPS[[mech]])) {
    seed <- SEED0 + r
    for (v in TARGET) {
      partner <- setdiff(TARGET, v)
      M  <- .amputate(num, v, rate, mech, partner = partner, seed = seed)
      Dm <- num; Dm[M, v] <- NA
      truth <- num[[v]]
      for (meth in METHODS_ORIG) {
        imp <- impute_one(Dm, v, meth, seed)
        rows[[k]] <- data.frame(
          mechanism = mech, rate = rate, seed = seed,
          variable = v, method = meth,
          nrmse   = nrmse(imp, truth, M),
          mae     = mae_metric(imp, truth, M),
          iqrnrmse = iqr_nrmse(imp, truth, M),
          stringsAsFactors = FALSE)
        k <- k + 1L
      }
    }
  }
}
res <- do.call(rbind, rows)

cat("\n=== SECTION 2: TABLE 2: NRMSE distribution (median; SD; max) ===\n")
for (v in TARGET) for (mech in c("MCAR","MAR","MNAR")) {
  for (meth in METHODS_ORIG) {
    x <- res$nrmse[res$variable==v & res$mechanism==mech & res$method==meth]
    cat(sprintf("%-5s %-12s %-26s md=%6.2f SD=%7.2f max=%7.1f\n",
                mech, meth, v, median(x), sd(x), max(x)))
  }
}

## ======================================================================
## SECTION 3: Sec 4.3 — tail-recovery under MCAR vs MNAR
## ======================================================================
cat("\n=== SECTION 3: SEC 4.3: tail-recovery (HH Aide FTE, missForest, >=P90) ===\n")
for (mech in c("MCAR","MNAR")) {
  rec <- c(); blw <- c()
  for (r in seq_len(10L)) {
    seed <- SEED0 + r
    M    <- .amputate(num, HHAIDE, 0.20, mech, partner = DIRECT, seed = seed)
    Dm   <- num; Dm[M, HHAIDE] <- NA
    set.seed(seed)
    imp  <- missForest(Dm, ntree = NTREE, verbose = FALSE)$ximp[[HHAIDE]]
    tr   <- tail_recovery(imp, num[[HHAIDE]], M, q = 0.90)
    rec  <- c(rec, tr["recovered"]); blw <- c(blw, tr["frac_below_half"])
  }
  cat(sprintf("  %-5s  magnitude recovered = %3.0f%% ; fraction below half true = %3.0f%%\n",
              mech, 100*mean(rec, na.rm=TRUE), 100*mean(blw, na.rm=TRUE)))
}

## ======================================================================
## SECTION 4: Sec 4.4 — co-deletion + exact binomial test
## ======================================================================
cat("\n=== SECTION 4: SEC 4.4: co-deletion + exact binomial test ===\n")
for (mech in c("MCAR","MAR","MNAR")) {
  overlap <- 0L; ndel <- 0L
  for (rate in RATES) for (r in seq_len(REPS[[mech]])) {
    seed <- SEED0 + r
    MA   <- .amputate(num, DIRECT, rate, mech, partner = HHAIDE, seed = seed)
    MB   <- .amputate(num, HHAIDE, rate, mech, partner = DIRECT, seed = seed)
    overlap <- overlap + sum(MA & MB); ndel <- ndel + sum(MA)
  }
  p_chance <- mean(RATES)
  bt <- binom.test(overlap, ndel, p = p_chance, alternative = "greater")
  cat(sprintf("  %-5s co-deletion = %2.0f%% (%d/%d) ; chance = %2.0f%% ; binomial p = %.3g\n",
              mech, 100*overlap/ndel, overlap, ndel, 100*p_chance, bt$p.value))
}

## ======================================================================
## SECTION 5: Sec 4.1 — threshold robustness
## ======================================================================
cat("\n=== SECTION 5: SEC 4.1: threshold robustness ===\n")
thr_grid <- expand.grid(sk = c(2, 2.5, 3, 5, 10), rr = c(0.80, 0.85, 0.90, 0.95))
for (i in seq_len(nrow(thr_grid))) {
  p  <- grip_profile(num, skew_thr = thr_grid$sk[i], collinearity_thr = thr_grid$rr[i])
  fl <- p$variable[p$at_risk]
  cat(sprintf("  skew>=%-4g |r|>=%.2f : %d flagged {%s}\n",
              thr_grid$sk[i], thr_grid$rr[i], length(fl), paste(fl, collapse=", ")))
}

## ======================================================================
## SECTION 6: Sec 4.5 — bootstrap five-replication mean
## ======================================================================
cat("\n=== SECTION 6: SEC 4.5: bootstrap five-replication mean (Direct, MCAR) ===\n")
dir_mcar <- res$nrmse[res$variable==DIRECT & res$mechanism=="MCAR" &
                      res$method=="missForest"]
set.seed(42)
boot <- replicate(10000, mean(sample(dir_mcar, 5, replace = TRUE)))
cat(sprintf("  median 5-rep mean = %.1f ; P(mean>=16.9) = %.0f%% ; P(single>=16.9) = %.0f%%\n",
            median(boot), 100*mean(boot>=16.9), 100*mean(dir_mcar>=16.9)))
for (mech in c("MCAR","MAR","MNAR")) {
  dD <- res$nrmse[res$variable==DIRECT & res$mechanism==mech & res$method=="missForest"]
  dH <- res$nrmse[res$variable==HHAIDE & res$mechanism==mech & res$method=="missForest"]
  n  <- min(length(dD), length(dH))
  cat(sprintf("  Ordering reversal %-5s : %.0f%%\n", mech,
              100*mean(dH[seq_len(n)] > dD[seq_len(n)])))
}

## ======================================================================
## SECTION 7: Table 3 — illustrative five-replication run
## ======================================================================
cat("\n=== SECTION 7: TABLE 3: illustrative five-replication run ===\n")
t3 <- data.frame()
for (s in 1:5) {
  Ma <- .amputate(num, DIRECT, 0.20, "MCAR", seed = s)
  Mh <- .amputate(num, HHAIDE, 0.20, "MCAR", seed = s)
  Da <- num; Da[Ma, DIRECT] <- NA
  Dh <- num; Dh[Mh, HHAIDE] <- NA
  set.seed(s); iD <- missForest(Da, ntree = NTREE, verbose = FALSE)$ximp[[DIRECT]]
  set.seed(s); iH <- missForest(Dh, ntree = NTREE, verbose = FALSE)$ximp[[HHAIDE]]
  t3 <- rbind(t3, data.frame(
    replication   = s,
    Direct_NRMSE  = round(nrmse(iD, num[[DIRECT]], Ma), 2),
    HHAide_NRMSE  = round(nrmse(iH, num[[HHAIDE]], Mh), 2)))
}
print(t3, row.names = FALSE)
cat(sprintf("  Direct mean %.1f (SD %.1f) ; HH mean %.1f (SD %.1f)\n",
            mean(t3$Direct_NRMSE), sd(t3$Direct_NRMSE),
            mean(t3$HHAide_NRMSE), sd(t3$HHAide_NRMSE)))

## ======================================================================
## SECTION 8: Sec 4.7 — OUTLIER SENSITIVITY [NEW]
## Remove top 1% of HH Aide FTE and re-run GRIP Steps 1 and 2
## ======================================================================
cat("\n=== SECTION 8: SEC 4.7: OUTLIER SENSITIVITY (top 1% removed) ===\n")

thr_99  <- quantile(num[[HHAIDE]], 0.99)
n_above <- sum(num[[HHAIDE]] > thr_99)
num_trimmed <- num[num[[HHAIDE]] <= thr_99, , drop = FALSE]
cat(sprintf("  Removing %d agencies above 99th percentile (HH Aide FTE > %.1f)\n",
            n_above, thr_99))
cat(sprintf("  Trimmed dataset: %d agencies\n", nrow(num_trimmed)))

## Step 1 on trimmed data
prof_trim <- grip_profile(num_trimmed, skew_thr = 3, collinearity_thr = 0.9)
cat("  Step 1 (trimmed):\n")
print(prof_trim[prof_trim$at_risk | prof_trim$variable %in% TARGET, ],
      row.names = FALSE)

## Step 2 (MCAR only, 10 reps) on trimmed data
cat("  Step 2 (MCAR, 10 reps, missForest, trimmed):\n")
for (v in TARGET) {
  nrmses_trim <- numeric(10L)
  for (r in seq_len(10L)) {
    seed <- SEED0 + r
    partner <- setdiff(TARGET, v)
    M  <- .amputate(num_trimmed, v, 0.20, "MCAR", partner = partner, seed = seed)
    Dm <- num_trimmed; Dm[M, v] <- NA
    set.seed(seed)
    imp <- missForest(Dm, ntree = NTREE, verbose = FALSE)$ximp[[v]]
    nrmses_trim[r] <- nrmse(imp, num_trimmed[[v]], M)
  }
  cat(sprintf("  %-26s MCAR SD = %.2f (cf. %.2f full data)\n",
              v, sd(nrmses_trim),
              sd(res$nrmse[res$variable==v & res$mechanism=="MCAR" &
                           res$method=="missForest"])))
}

## ======================================================================
## SECTION 9: MICE(PMM) rows for Table 2 [NEW]
## ======================================================================
cat("\n=== SECTION 9: TABLE 2 MICE(PMM) supplementary rows ===\n")
cat("  NOTE: MICE(PMM) is computationally intensive.\n")
cat("  Using 30 MCAR reps, 30 MAR reps, 10 MNAR reps at 20% rate only.\n")
cat("  Full multi-rate runs: set full_mice = TRUE below.\n")

full_mice <- FALSE    ## set TRUE to run all rates (very slow)
mice_rates <- if (full_mice) RATES else 0.20

rows_mice <- list(); km <- 1L
for (mech in c("MCAR", "MAR", "MNAR")) {
  n_reps_mice <- REPS[[mech]]
  for (rate in mice_rates) for (r in seq_len(n_reps_mice)) {
    seed <- SEED0 + r
    for (v in TARGET) {
      partner <- setdiff(TARGET, v)
      M  <- .amputate(num, v, rate, mech, partner = partner, seed = seed)
      Dm <- num; Dm[M, v] <- NA
      imp <- impute_one(Dm, v, "mice_pmm", seed)
      rows_mice[[km]] <- data.frame(
        mechanism = mech, rate = rate, seed = seed,
        variable = v, method = "mice_pmm",
        nrmse    = nrmse(imp, num[[v]], M),
        stringsAsFactors = FALSE)
      km <- km + 1L
    }
  }
}
res_mice <- do.call(rbind, rows_mice)

cat("\n  MICE(PMM) results:\n")
for (v in TARGET) for (mech in c("MCAR","MAR","MNAR")) {
  x <- res_mice$nrmse[res_mice$variable==v & res_mice$mechanism==mech]
  cat(sprintf("  %-5s mice_pmm %-26s md=%6.2f SD=%7.2f max=%7.1f\n",
              mech, v, median(x), sd(x), max(x)))
}

## ======================================================================
## SECTION 10: SUPPLEMENTAL TABLE 1 — MAE and IQR-NRMSE [NEW]
## ======================================================================
cat("\n=== SECTION 10: SUPPLEMENTAL TABLE 1: MAE and IQR-NRMSE ===\n")
cat("  Alternative metrics for the two at-risk variables, missForest, MCAR:\n")
for (v in TARGET) {
  mae_vals  <- res$mae[res$variable==v & res$mechanism=="MCAR" & res$method=="missForest"]
  iqr_vals  <- res$iqrnrmse[res$variable==v & res$mechanism=="MCAR" & res$method=="missForest"]
  nrm_vals  <- res$nrmse[res$variable==v & res$mechanism=="MCAR" & res$method=="missForest"]
  cat(sprintf("\n  %s\n", v))
  cat(sprintf("    NRMSE    : med=%.2f SD=%.2f max=%.1f\n",
              median(nrm_vals), sd(nrm_vals), max(nrm_vals)))
  cat(sprintf("    IQR-NRMSE: med=%.2f SD=%.2f max=%.1f\n",
              median(iqr_vals, na.rm=TRUE), sd(iqr_vals, na.rm=TRUE), max(iqr_vals, na.rm=TRUE)))
  cat(sprintf("    MAE      : med=%.1f SD=%.1f max=%.0f\n",
              median(mae_vals), sd(mae_vals), max(mae_vals)))
}

cat("\nAll analyses complete.\n")
```
