## Additional File 4_Code_3_Grid for "Geometry-Aware Reproducibility of Imputation Protocol (GRIP): diagnosing two failure modes of single imputation for heavy-tailed, collinear variables in biomedical data": Additional File 4_Code_3_Grid.R

```
########################################################################
##  Supplemental Code 3 — Synthetic Grid Simulation with missForest
##
##  Purpose: Map the LOUD/SILENT/STABLE failure-mode boundary across the
##  (skewness, Pearson |r|) parameter space for non-extrapolating imputers.
##  Complements Supplemental Code 2 (CMS demonstration) and the OLS-based
##  grid reported in Table 4 of the main manuscript.
##
##  Design follows ADEMP (Morris, White & Crowther, Stat Med 2019):
##    Aims     : Characterize LOUD and SILENT failure rates across
##               (sigma, rho) combinations; overlay CMS variables.
##    DGP      : Bivariate lognormal via Gaussian copula (n = 300).
##               X1 = exp(sigma * Z1), X2 = exp(sigma * Z2),
##               (Z1, Z2) ~ N(0, [[1, rho],[rho, 1]]).
##               Five independent N(0,1) nuisance variables.
##    Estimands: SD(NRMSE) under MCAR (LOUD index);
##               mean tail-recovery under MNAR (SILENT index);
##               co-deletion rate under MNAR (mechanism index).
##    Methods  : missForest (ntree = 20; reduced for speed; set NTREE=100
##               for publication-quality results); mean imputation.
##    Performance: LOUD if SD(NRMSE) >= 1.0; SILENT if tail-recovery < 50%.
##
##  Computation note: Full grid (20 cells × 2 mechanisms × REPS) with
##  NTREE = 100 takes approximately 4–8 hours on a single core. Set
##  NTREE = 20 for a screening run (30–60 min). Results are qualitatively
##  identical; NRMSE values differ slightly.
##
##  Output: grid_results data frame + phase diagram plot (PNG).
##
##  Author : Claire Su-Yeon Park (ORCID 0000-0002-2109-9885)
##  License: MIT
##  Dependencies: install.packages(c("missForest", "e1071"))
########################################################################

suppressMessages({ library(missForest); library(e1071) })
source("Supplemental_Code_1_GRIP.R")   # nrmse(), tail_recovery(), .amputate()

## ── user-adjustable parameters ────────────────────────────────────────────
N       <- 300L          ## cases per simulated dataset
NTREE   <- 20L           ## trees (set 100 for publication)
N_MCAR  <- 20L           ## MCAR replications per cell
N_MNAR  <- 10L           ## MNAR replications per cell
RATE    <- 0.20          ## missingness rate
SEED0   <- 2000L
OUT_DIR <- "."           ## directory for output PNG

## ── parameter grid ────────────────────────────────────────────────────────
## sigma levels: lognormal sigma -> approximate skewness
##   sigma=0.3 -> skew ~0.9
##   sigma=0.7 -> skew ~3
##   sigma=1.1 -> skew ~8
##   sigma=1.5 -> skew ~35
SIGMA_LEVELS <- c(0.3, 0.7, 1.1, 1.5)
RHO_LEVELS   <- c(0.50, 0.70, 0.85, 0.95, 0.99)

## ── helper: sample skewness ───────────────────────────────────────────────
skew_s <- function(x) {
  x <- x[!is.na(x)]; m <- mean(x)
  mean((x - m)^3) / (mean((x - m)^2)^1.5)
}

## ── main loop ─────────────────────────────────────────────────────────────
grid_rows <- list(); row_idx <- 1L

for (sigma in SIGMA_LEVELS) {
  for (rho in RHO_LEVELS) {

    set.seed(as.integer((sigma * 10 + rho * 100) * 7))
    Z1  <- rnorm(N)
    Z2  <- rho * Z1 + sqrt(1 - rho^2) * rnorm(N)
    X1  <- exp(sigma * Z1)
    X2  <- exp(sigma * Z2)
    nuis <- matrix(rnorm(N * 5L), N, 5L)
    D   <- as.data.frame(cbind(X1, X2, nuis))
    names(D) <- c("v1", "v2", paste0("n", 1:5))

    actual_skew <- skew_s(X1)
    actual_r    <- cor(X1, X2)

    ## ── MCAR stress test ─────────────────────────────────────────────────
    mcar_nrmse <- numeric(N_MCAR)
    for (r in seq_len(N_MCAR)) {
      seed <- SEED0 + r
      M    <- .amputate(D, "v1", RATE, "MCAR", partner = "v2", seed = seed)
      Dm   <- D; Dm[M, "v1"] <- NA
      set.seed(seed)
      tryCatch({
        imp <- missForest(Dm, ntree = NTREE, verbose = FALSE)$ximp[["v1"]]
        mcar_nrmse[r] <- nrmse(imp, D[["v1"]], M)
      }, error = function(e) mcar_nrmse[r] <<- NA_real_)
    }

    ## ── MNAR stress test + tail recovery ─────────────────────────────────
    mnar_nrmse   <- numeric(N_MNAR)
    mnar_tailrec <- numeric(N_MNAR)
    co_del_rate  <- numeric(N_MNAR)
    q90          <- quantile(D[["v1"]], 0.90)
    q90_v2       <- quantile(D[["v2"]], 0.90)

    for (r in seq_len(N_MNAR)) {
      seed <- SEED0 + r * 3L
      M1   <- .amputate(D, "v1", RATE, "MNAR", partner = "v2", seed = seed)
      M2   <- .amputate(D, "v2", RATE, "MNAR", partner = "v1", seed = seed)
      Dm   <- D; Dm[M1, "v1"] <- NA
      set.seed(seed)
      tryCatch({
        imp <- missForest(Dm, ntree = NTREE, verbose = FALSE)$ximp[["v1"]]
        mnar_nrmse[r] <- nrmse(imp, D[["v1"]], M1)
        tr <- tail_recovery(imp, D[["v1"]], M1, q = 0.90)
        mnar_tailrec[r] <- tr["recovered"]
        ## Co-deletion: extreme X1 deleted AND extreme X2 also deleted
        ext1_del <- M1 & (D[["v1"]] >= q90)
        if (any(ext1_del))
          co_del_rate[r] <- mean(M2[ext1_del])
        else
          co_del_rate[r] <- NA_real_
      }, error = function(e) {
        mnar_nrmse[r]   <<- NA_real_
        mnar_tailrec[r] <<- NA_real_
        co_del_rate[r]  <<- NA_real_
      })
    }

    mcar_sd    <- sd(mcar_nrmse, na.rm = TRUE)
    mnar_trec  <- mean(mnar_tailrec, na.rm = TRUE)
    mnar_codel <- mean(co_del_rate, na.rm = TRUE)

    mode_mcar  <- if (!is.na(mcar_sd) && mcar_sd >= 1.0) "LOUD" else "STABLE"
    mode_mnar  <- if (!is.na(mnar_trec) && mnar_trec < 0.50 &&
                       sd(mnar_nrmse, na.rm=TRUE) < 0.50) "SILENT" else "STABLE"

    grid_rows[[row_idx]] <- data.frame(
      sigma        = sigma,
      rho_copula   = rho,
      actual_skew  = round(actual_skew, 1),
      actual_r     = round(actual_r, 3),
      mcar_sd      = round(mcar_sd, 3),
      mcar_med     = round(median(mcar_nrmse, na.rm=TRUE), 3),
      mnar_sd      = round(sd(mnar_nrmse, na.rm=TRUE), 3),
      mnar_med     = round(median(mnar_nrmse, na.rm=TRUE), 3),
      tail_rec_pct = round(mnar_trec * 100, 1),
      co_del_pct   = round(mnar_codel * 100, 1),
      mode_mcar    = mode_mcar,
      mode_mnar    = mode_mnar,
      stringsAsFactors = FALSE
    )

    cat(sprintf("sigma=%.1f rho=%.2f | skew=%5.1f r=%5.3f | "
                "MCAR SD=%6.3f (%s) | "
                "MNAR tailrec=%4.0f%% codel=%3.0f%% (%s)\n",
                sigma, rho, actual_skew, actual_r,
                mcar_sd, mode_mcar,
                mnar_trec * 100, mnar_codel * 100, mode_mnar))

    row_idx <- row_idx + 1L
  }
}

grid_results <- do.call(rbind, grid_rows)

## ── print summary table ───────────────────────────────────────────────────
cat("\n", strrep("=", 110), "\n")
cat(sprintf(
  "%-6s %-6s %6s %6s | %8s %8s %10s | %7s %7s %6s %7s %10s\n",
  "sigma", "rho", "Skew", "|r|",
  "MCAR_SD", "MCAR_med", "Mode_MCAR",
  "MNAR_SD", "MNAR_med", "TailRec", "Codel%", "Mode_MNAR"))
for (i in seq_len(nrow(grid_results))) {
  r <- grid_results[i, ]
  cat(sprintf(
    "%-6.1f %-6.2f %6.1f %6.3f | %8.3f %8.3f %10s | %7.3f %7.3f %6.1f %7.1f %10s\n",
    r$sigma, r$rho_copula, r$actual_skew, r$actual_r,
    r$mcar_sd, r$mcar_med, r$mode_mcar,
    r$mnar_sd, r$mnar_med, r$tail_rec_pct, r$co_del_pct, r$mode_mnar))
}

## ── phase diagram plot ────────────────────────────────────────────────────
png(file.path(OUT_DIR, "GRIP_phase_diagram.png"),
    width = 900, height = 700, res = 120)
par(mfrow = c(1, 2), mar = c(4.5, 4.5, 3, 1))

## Panel A: MCAR failure mode (LOUD indicator = SD)
cols_mcar <- ifelse(grid_results$mode_mcar == "LOUD",   "#CC0000",
             ifelse(grid_results$mode_mcar == "STABLE", "#2166AC", "#888888"))
plot(grid_results$actual_skew, grid_results$actual_r,
     pch = 21, bg = cols_mcar, cex = 2.2,
     xlim = c(0, 45), ylim = c(0, 1),
     xlab = "Sample skewness", ylab = "Pearson |r|",
     main = "A. MCAR failure mode (missForest)")
abline(v = 3, lty = 2, col = "gray40"); abline(h = 0.9, lty = 2, col = "gray40")
legend("bottomright", legend = c("LOUD", "STABLE"),
       pt.bg = c("#CC0000","#2166AC"), pch = 21, pt.cex = 1.5, bty = "n")
## Overlay CMS variables
points(c(38.6, 43.3), c(0.96, 0.96), pch = 4, cex = 2.5, lwd = 2, col = "black")
text(c(38.6, 43.3), c(0.93, 0.93), c("Dir.","HH"), cex = 0.7)

## Panel B: MNAR failure mode (SILENT indicator = tail recovery)
cols_mnar <- ifelse(grid_results$mode_mnar == "SILENT", "#E08214",
             ifelse(grid_results$mode_mnar == "STABLE",  "#2166AC", "#888888"))
plot(grid_results$actual_skew, grid_results$actual_r,
     pch = 21, bg = cols_mnar, cex = 2.2,
     xlim = c(0, 45), ylim = c(0, 1),
     xlab = "Sample skewness", ylab = "Pearson |r|",
     main = "B. MNAR failure mode (missForest)")
abline(v = 3, lty = 2, col = "gray40"); abline(h = 0.9, lty = 2, col = "gray40")
legend("bottomright", legend = c("SILENT (tail-loss)", "STABLE"),
       pt.bg = c("#E08214","#2166AC"), pch = 21, pt.cex = 1.5, bty = "n")
points(c(38.6, 43.3), c(0.96, 0.96), pch = 4, cex = 2.5, lwd = 2, col = "black")
text(c(38.6, 43.3), c(0.93, 0.93), c("Dir.","HH"), cex = 0.7)

dev.off()
cat("\nPhase diagram written to: GRIP_phase_diagram.png\n")
cat("Grid results object: grid_results\n")
cat("Done.\n")
```
